# Experiences and impacts of side effects among contraceptive users in the UK: exploring individual narratives of contraceptive side effects

**DOI:** 10.1101/2023.10.02.23296334

**Authors:** Catherine Stewart, Rose Stevens, Fiona Kennedy, Paulina Cecula, Elena Rueda Carrasco, Jennifer Hall

## Abstract

**Purpose:** While many women worldwide use contraception, there is a paucity of research on individual experiences of side effects and their impacts. To address this gap, we analysed free-text responses of contraception experiences from 337 women aged 18 to 35, based in the UK who took part in an online survey on contraception.

**Materials and methods:** Through a directed content analysis approach, we developed a coding framework based on existing literature and initial response review. It included six themes; method(s) of contraception, side effect(s) experienced, impact of side effect(s), timing of side effect(s), interactions with healthcare practitioners, and trial and error.

**Results:** Side effect experiences and impacts varied greatly between individuals and contraceptives. Most participants described negative effects, such as mental health issues and bleeding problems. Some shared positive experiences related to bleeding management and the absence of side effects. Some experienced side effects after years of use and felt unheard by practitioners.

**Conclusions:** This contraceptive experience variability underscores the need for further research into individual side effect variation. We advocate for a patient-centred approach to contraceptive counselling. Practitioners should play an active role in improving contraception prescription, acknowledging the diverse experiences and preferences of patients.

## Introduction

Globally, there are roughly 842 million women of reproductive age using modern contraceptive methods [1]. Since their development, hormonal forms of contraception (such as the combined pill, progestogen-only pill, intrauterine system, implant, and injection) have seen relatively high effectiveness for their original intended use; pregnancy prevention [2]. Women commonly use these medications for around three quarters of their reproductive lives, both for pregnancy prevention and other uses such as bleeding control and acne reduction. We note that not all users of hormonal contraception identify as women and that not all women will want or need to use contraception.

As well as the intended effects, many users of contraception report experiencing unintended effects from contraception (here referred to as “side effects”). About one-third of US contraceptive users (31%) say they are experiencing side effects from their current method, with just over half (52%) reporting the side effects to be more severe than expected [3]. Whilst side effects are not by definition always negative, contraceptive side effects can cause dissatisfaction with and subsequent discontinuation of a method, increasing the risk of unintended pregnancy [4,5]. Additionally, a recent systematic review shows that the desire to avoid negative side effects was the primary reason for rejecting hormonal contraception in Western countries, regardless of whether the individual had experienced negative effects themselves [6]. The impact of side effects and how they influence contraceptive decisions are heavily contingent upon women’s education, lifestyle, cultural beliefs, partner and peer influence, as well as health practitioners’ behaviours, skills and availability and thus vary greatly by the context in which a contraceptive is used [7].

The aim of this study was to explore the experiences and impacts of contraceptive side effects on women in the UK.

## Methods

### Study setting and design

Dama Health is a women’s health start-up based in the UK. The Dama Health Genetics and Contraception Study (DHGCS) sought to explore variation in the experience of contraceptive side effects that led to discontinuation of a contraceptive method, using both genetic and survey data.

The DHGCS was a case-control study administered from mid-October 2022 through April 2023. It utilised a mixed-methods design, gathering online survey data on participants’ demographics, medical history, symptoms and contraceptive experiences and genetic data from self-administered saliva samples. The survey was hosted on a secure platform, Qualtrics XM. Participants could complete the study at a convenient time and place.

### Participants

Eligibility criteria were 18 to 35 years of age, assigned female at birth, and residing in the UK at the time of the study. Cases were participants who had experienced one or more severe side effects from the combined pill, progestogen-only pill, contraceptive implant, or intrauterine hormone-releasing system (within the past 10 years) which led to discontinuation of that method. Controls were participants who were using any of these contraceptive methods without experiencing any side effects (within the past 10 years). Participants self-selected as either case or control. Electronic informed consent was given.

Based on funding constraints for the genetic arm, we recruited 1,135 participants through convenience sampling via social media advertisements.

At the end of the survey, participants in the case group were asked ‘Is there anything else that you’d like to share with us about the contraception you’ve tried in the past, your experience with it and side effects?’ Participants in the control group were asked ‘Is there anything else that you’d like to share with us about the contraception you’ve tried in the past and your experience with it?’

### Data analysis

We took a directed content analysis approach, developing a coding framework based on background knowledge and existing literature on the topic which was augmented by inductive codes from an initial review of responses and team discussion [8]. This deductive style of coding fits well to less in-depth data and allows for descriptive insights into types of content most mentioned as well homing in on particular contraceptive experiences during analysis. Data were managed using Excel. The final coding framework included six themes: method(s) of contraception, side effect(s) experienced (both negative and positive), impact of side effect(s), timing of side effect(s), interactions with healthcare practitioners, and trial and error.

Coding and analysis was led by CS, a white, female researcher, with an MSc and qualitative research experience. CS was supervised by JH, a female medical doctor and Public Health researcher with training and experience of qualitative research. RS is a white, female mixed-methods doctoral researcher. Codes, the coding framework and findings were regularly discussed with the wider team.

### Ethical approval

The DHGCS was given a favourable ethical opinion by the Reading Independent Ethics Committee (UK) on October 14, 2022.

## Results

A total of 1,135 participants took part in the pseudonymised online survey; 796 cases and 337 controls. Two participants withdrew after completion without providing a reason. Data were retained only for participants who completed the whole survey.

Overall, 337 participants (30%) provided free-text responses; 262 from the case group (33%) and 75 from the control group (22%). Socio-demographics are shown in Table 1. Those providing free-text responses were slightly older and more had ever been pregnant but were otherwise similar to the whole sample.

**Table 1.**
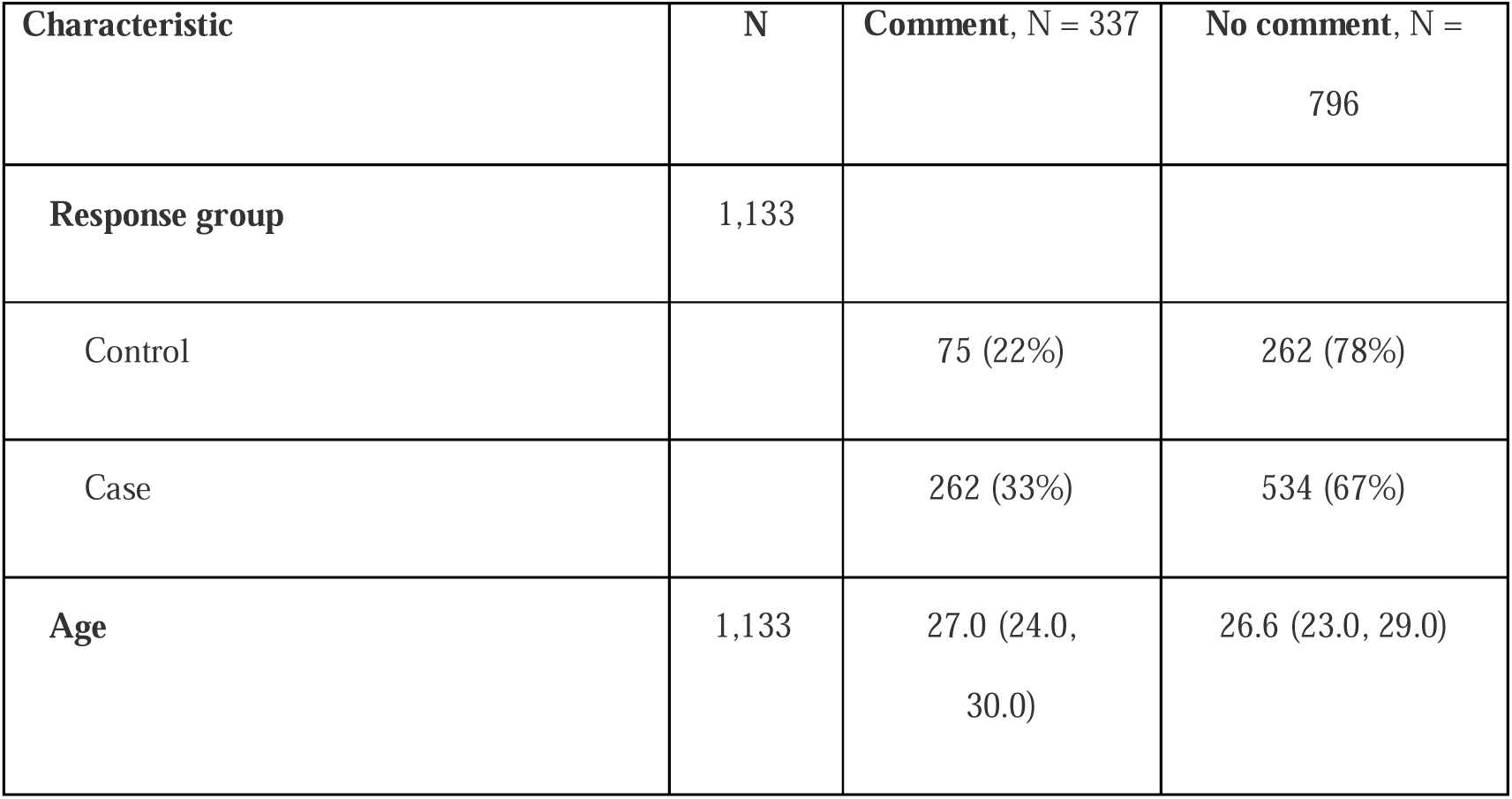
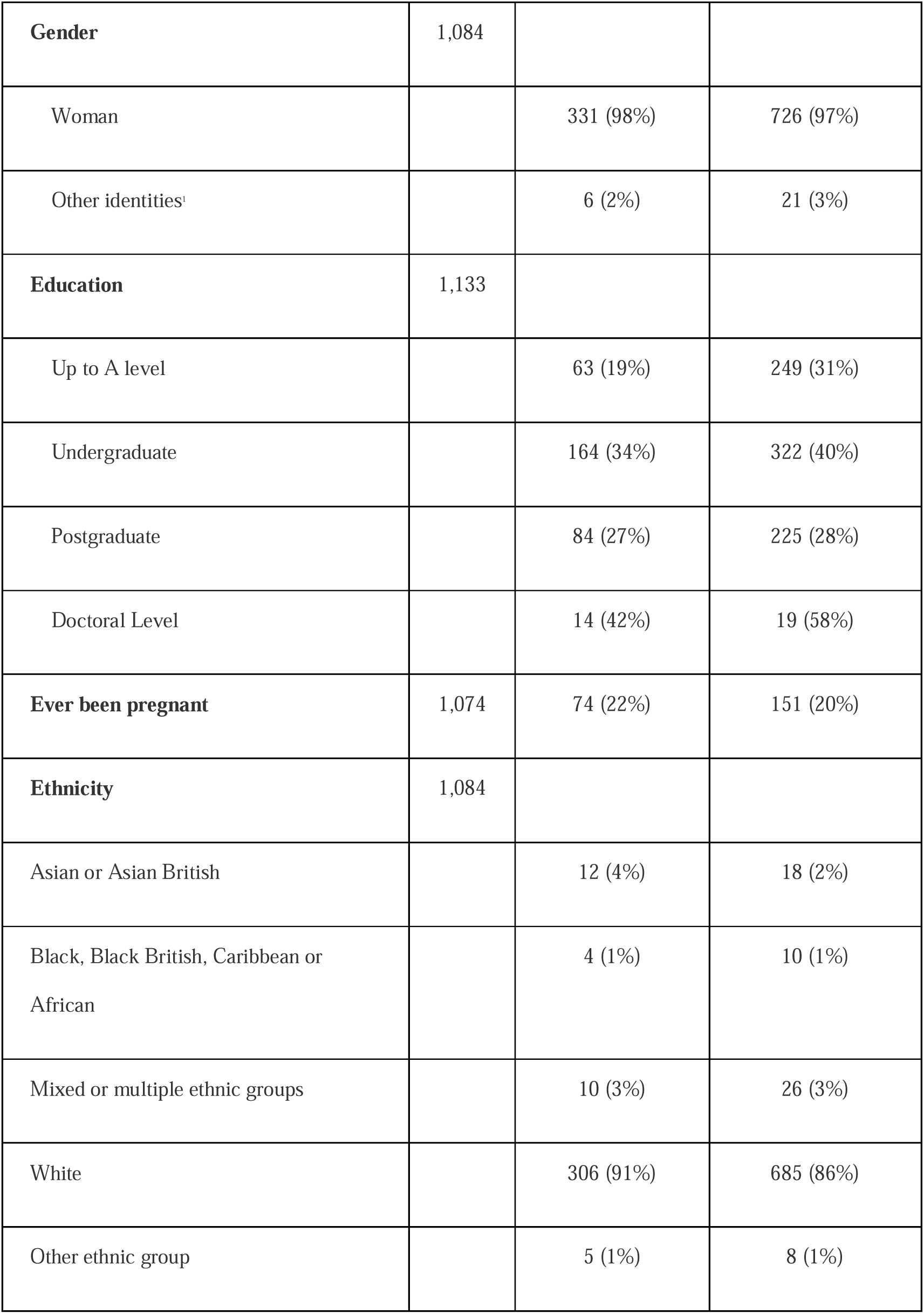
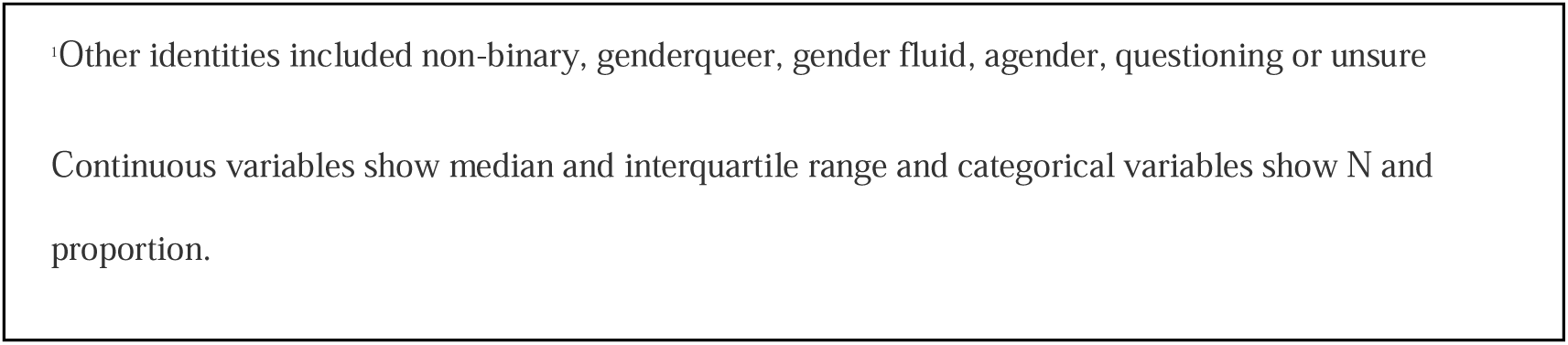
Sociodemographics of all participants who commented and did not comment on the free-text box.

| Characteristic | N | Comment, N = 337 | No comment, N = 796 |
| --- | --- | --- | --- |
| Response group | 1,133 |  |  |
| Control |  | 75 (22%) | 262 (78%) |
| Case |  | 262 (33%) | 534 (67%) |
| Age | 1,133 | 27.0 (24.0, 30.0) | 26.6 (23.0, 29.0) |
| **Gender** | 1,084 |  |  |
| Woman |  | 331 (98%) | 726 (97%) |
| Other identities<sup>1</sup> |  | 6 (2%) | 21 (3%) |
| **Education** | 1,133 |  |  |
| Up to A level |  | 63 (19%) | 249 (31%) |
| Undergraduate |  | 164 (34%) | 322 (40%) |
| Postgraduate |  | 84 (27%) | 225 (28%) |
| Doctoral Level |  | 14 (42%) | 19 (58%) |
| **Ever been pregnant** | 1,074 | 74 (22%) | 151 (20%) |
| **Ethnicity** | 1,084 |  |  |
| Asian or Asian British |  | 12 (4%) | 18 (2%) |
| Black, Black British, Caribbean or African |  | 4 (1%) | 10 (1%) |
| Mixed or multiple ethnic groups |  | 10 (3%) | 26 (3%) |
| White |  | 306 (91%) | 685 (86%) |
| Other ethnic group |  | 5 (1%) | 8 (1%) |

Other identities included non-binary, genderqueer, gender fluid, agender, questioning or unsure
Continuous variables show median and interquartile range and categorical variables show N and proportion.

### Method of contraception

All methods of hormonal contraception considered in this study were mentioned by participants, with the contraceptive pill (n=190) mentioned most commonly (Table 2). Ten participants discussed using multiple methods simultaneously; notably being prescribed the contraceptive pill while on the implant for symptom control.

**Table 2.**
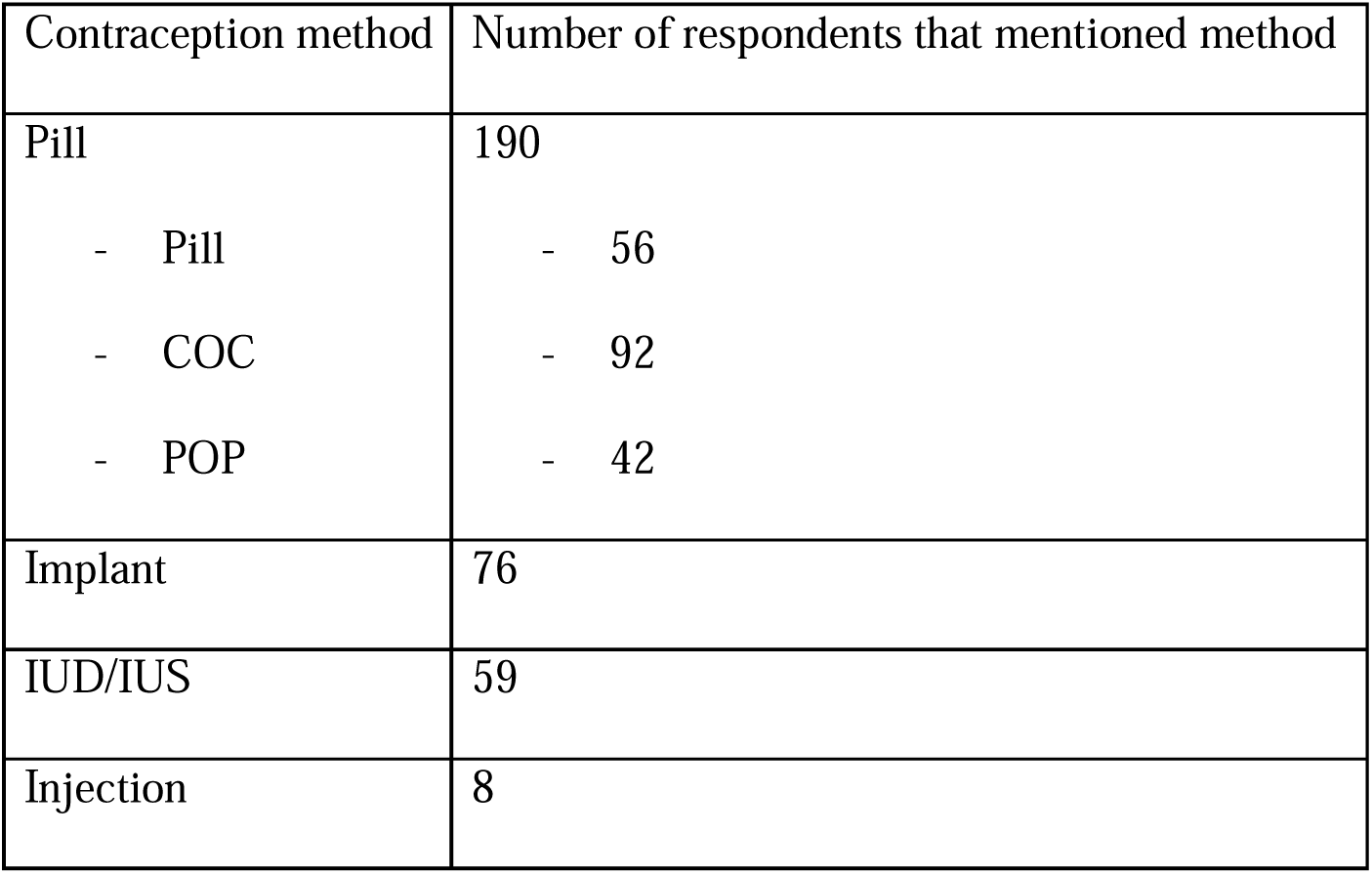
Contraception method by number of respondents that mentioned.

| Contraception method | Number of respondents that mentioned method |
| --- | --- |
| Pill | 190 |
| - Pill | - 56 |
| - COC | - 92 |
| - POP | - 42 |
| Implant | 76 |
| IUD/IUS | 59 |
| Injection | 8 |

### Side effects

Seventy-five percent (n=253) of all participants who responded to the free-text box described experiencing at least one side effect from at least one contraceptive method. While the majority of participants described negative side effects (n=205), positive side effects were also discussed (n=46).

The main negative side effects mentioned were a) mental health including mood issues, depression, anxiety and feeling suicidal (n=82); “Rigevidon made me feel like a raging, depressed, anxious monster constantly” (COC, 20-24 years, Woman, White British, Postgraduate, Case), b) menstrual bleeding (n=78), including constant bleeding/spotting (n=46), irregular (n=20), painful or heavy bleeding (n=12)); “various different side effects with bleeding-either a lot, spotting, or painful” (COC/POP, 20-24 years, Woman, White British, Postgraduate, Case), and c) headaches/migraines (n=34); “severe migraines multiple times a week that would last hours” (COC, 25-39 years, Woman, White British, Postgraduate, Case).

The main positive side effects discussed were a) stopping/significantly reducing bleeding (n=21); “The main benefit…of the coil for me is stopping my periods” (IUS/IUD, 30-34 years, Woman, Indian, Postgraduate, Control), and b) reducing the heaviness or painfulness of bleeding (n=18). Experiencing no side effects was also commonly discussed in a positive way (n=44), e.g. “I am doing very well on the minipill (Cerazette) now with no side effects” (POP, 20-24 years, Woman, Chinese, Undergraduate, Case).

There were associations between different methods and side effects. For example, the implant was commonly associated with constant bleeding/spotting (n=26 out of 60 symptom reports on the implant). The pill was more commonly associated with mental health concerns (n=44/132) and, specifically for COC, headaches (n=27/57). There was a great deal of variation in experiences, with opposing side effects described by different participants for the same method of contraception.

### Impact of side effects

30 participants discussed the impact of their side effects on their relationships, sex life, work/education or mental health; 24 negatively and six positively. Negative impacts on their relationship, included being “sick of feeling dirty from all the bleeding” (IUS/IUD, 30-34 years, Woman, White British, Postgraduate, Case) which led to a lack of intimacy, whereas those in whom the IUS resulted in no periods were positive about the impact on their sex lives, as well as not worrying about pregnancy. These impacts drove whether users considered using contraception to be a positive or negative experience overall. When the experience was positive, participants were likely to be on the same method for years, demonstrating the impact of side effect absence on continuity. For example, “I went onto the mini pill as my first form of contraception about 11 years ago and have been on it ever since, my periods stopped which for me was a huge bonus and haven’t seen any negative issues” (POP, 25-29 years, Woman, White British, Undergraduate, Control).

Descriptions of negative experiences were typically more detailed and used stronger language, conveying depth of feeling and the extent of the impact of the side effects. Participants used phrases such as “worst time of my life” (COC/POP, 20-24 years, Woman, White British, Postgraduate, Case), “horrible experience” (Implant, 25-29 years, Woman, White British, A-Levels, Case), and “life destroying” (Implant, 30-34 years, Woman, White British, Undergraduate, Case).

### Trial and error

Only one participant explicitly discussed the concept of “trial and error” but 32% (n=107) indicated switching method and/or brand of contraception at least once. Participants often described different experiences or side effects for each method they tried and for different brands of the same method. Many participants went on to ultimately find a method that suited them: “I… am finally happy after many years of trying contraception” (IUS/IUD, 25-29 years, Woman, Mixed, A-levels, Case).

### Timing of side effects

Participants described how their side effects had either varied throughout the course of using the contraception, appeared over time (often years after starting the contraception), or after insertion of a new LARC device (despite no previous side effects) (n=25). For example, “Some of the side effects only happened in the beginning and then resolved … some appeared with time, especially the ones related to mental health” (COC, 30-34 years, Other White, Postgraduate, Case).

### Health care interactions

Fourteen participants felt that healthcare practitioners had not believed the side effects they were experiencing, with participants saying that they were “ignored” (COC/POP, 20-25 years, Woman, White British, Undergraduate, Case), had “not been taken seriously” (POP, 30-34 years, Woman, White British, Undergraduate, Case), or that healthcare practitioners “wouldn’t believe it was the contraception causing the symptoms” (Implant/IUS/IUD, 20-24 years, Non-binary, White British, A-Levels, Case). Side effects starting sometime after the initiation of contraception were particularly likely to lead to negative healthcare interactions, including healthcare professionals acting as barriers to contraceptive removal.

Twelve participants described a lack of counselling about potential side effects and/or methods available. Participants felt health professionals were often lacking knowledge of contraception, and which methods might be best for them given their medical history. One participant said “Even though I explained to my doctor why I wanted the pill to control my PMDD [Premenstrual dysphoric disorder], it seems they did little research to find the best brand. I did my research and found that … Yasmin works best for PMDD… Since being put on Yasmin I have had limited side effects and can function properly - disappointed that my doctor wasn’t the one to understand this and save me the trauma” (COC, 20-24 years, Woman, Pakistani, Undergraduate, Case).

## Discussion

### Findings and interpretation

This study highlights how contraceptive experience is specific to the individual; side effects vary depending on the person, the method and the duration of use, as well as the significant impact that contraceptive side effects have on people’s relationships, work, education and mental health. Healthcare practitioners should consider this when assessing their patients’ needs.

Our findings suggest that individuals perceive side effects differently given the number of ‘control’ participants (those who self-selected as not experiencing side effects) who nevertheless discussed experiencing them. This may imply ambiguity surrounding what people classify as side effects; highlighting the difficulty in defining and measuring side effects quantitatively without information as to how they are perceived by users. It shows that people’s conceptualisation of side effects may be dynamic and rooted in personal preference. This nuance can be lost in communication between patients and practitioners when coming to a contraceptive decision [9]. Understanding these nuances is further limited by population health and clinical research studies which fail to capture comprehensive information on side effect experiences and patient-priorities due to a reliance on narrow researcher-driven measurement categories [10].

### Results in the Context of What is Known

The fact that our participants most commonly discussed the pill as a method and menstrual bleeding and mental health as side effects fits with prescribing rates and previous research on side effects, as does the common report of bleeding problems on the implant [11,12]. This study showed that discontinuation is often related to side effects, which is in line with data showing that side effects from contraceptive methods accounted for 38% of users changing methods [13]. The wide variation of experiences within the same methods of contraception supports research showing that side effect experiences, and their severity, vary considerably from person to person [14)]

Our study supports wider research showing the trade-off women face when they experience negative side effects, choosing between rejecting contraception altogether, switching between several methods until they find one that is acceptable to them, or enduring side effects and their impacts on their daily life and activities [15–18]. Conversely, contraception has also been linked to improving women’s quality of life in several domains [15,19,20], and our findings support research showing that positive side effects of contraception include improved sexual satisfaction through reduced anxiety and reduced bleeding [21]. This highlights the importance of finding the right method of contraception that suits the needs of the individual in the context of their wider lives and goals.

### Clinical Implications

The process of prescribing the best contraception for an individual is challenging; it requires time, knowledge of methods and their side effects, as well as an understanding that individuals react variably to each method and have different expectations or goals surrounding their contraceptive use. Healthcare workers may have additional and ongoing training needs in this regard. Within this study, participants felt they had not received sufficient counselling, and many felt that healthcare practitioners had not believed either the severity or cause of their symptoms and, consequently, had been unsupportive. This treatment brings into question a person’s contraceptive autonomy [22], as it may impact a person’s ability to make informed decisions or even restrict their choice to switch or discontinue their methods freely.

Taking a patient-centred approach to contraceptive counselling would ensure that the individual’s needs and preferences are taken into account [23], recognising the patient as the expert in their own experiences, values, and preferences [27]. Practitioners should aim to understand how side effect experiences and worries fit within a person’s wider life and explain the evidence available without dismissing users’ concerns [11]. To be able to counsel patients effectively, practitioners must know which contraceptive options are available, their potential side effects, and the typical risk of said side effects, especially among different groups of women. Even with extensive contraceptive counselling experience, a major role of the health practitioner may be to help manage patient expectations, educating patients about our current inability to predict who will experience which side effects or to what level from any given contraception. Consequently, the current prescription process for contraception is often trial-and-error. While not ideal, it is often successful, with many participants within this study going on to find a method of contraception they were happy with. Practitioners should stress that contraception should be re-discussed if symptoms with a severe impact, as determined by the patient, are experienced at any time so options can be re-evaluated. This may require a shift of mindset away from the classical efficacy-based approach of prescribing (for example, promotion of only LARCs) to an approach more weighted in patient acceptability [25].

### Research Implications

Our analysis highlights gaps in what users understand by ‘side effects’. Studies setting out to measure side effects over time using measurement tools informed by users’ priorities may help improve the information that can be given during counselling [12]. Future studies investigating the degree of interindividual variation in side effects and identifying predictors of different symptoms over time are also needed to support the development of precision medicine approaches to contraception, including decision support tools and biomarker tests, called for among participants in previous research [14,26,27]. These could provide personalised contraceptive recommendations to minimise the risk of side effects [28] and would respond to calls for more investment into improved contraceptive technologies [29] rather than ascribing to the idea that contraceptive side-effects are just ‘the price women pay’ for preventing pregnancy [14,30]. Finally, it would be important to examine the acceptability of the trial and error process and how it could be optimised to improve patient experience.

### Strengths and Limitations

The strength of this study lies within its ability to explore novel data from free-text responses from a large sample of contraceptive users of different methods. This overcomes limitations from both quantitative research, where narrow response categories may fail to capture the context or impact of contraceptive experience, and qualitative research, where small sample sizes and narrower scopes may cap the breadth of responses captured. A high proportion of participants chose to fill in the free-text box, at the end of a long survey (average length of 26 minutes), suggesting high participant engagement. This follows calls for collecting patient-reported outcomes and hearing patient priorities in their own words. It demonstrates women’s desire to engage in previously neglected areas of research, as evidenced by the response to the UK Government’s 2022 call for evidence regarding women’s health [31].

The main limitation is potential selection biases in recruitment and who answered the text box, limiting the representativeness of the data. By nature of the case and control design, extremes of experience are likely disproportionately represented and those most motivated to respond to the free-text box are also likely those with the strongest feelings about their experiences. Furthermore, the sample is highly educated and primarily white, limiting the generalisability of our conclusions.

## Conclusions

Our analysis revealed that, for most respondents, the path to finding an acceptable contraceptive is complex and unique. Many had strong sentiments about difficulties faced along this journey, reporting wide variation in side effects and the severity of their impacts. However, there was a strong perceived payoff if side-effect-free pregnancy protection was found. Respondents tried multiple methods, sought improved communication with health practitioners, and more information on personal side effect risks. Health practitioners’ ability to support women through this trial-and-error process may be limited by a lack of data on side effect experience variation and adequate training. Adopting patient-centred contraceptive counselling by valuing patients’ expertise and diversity in their experiences and preferences while maintaining expertise in the clinical information available may enable an improvement in the current standard of care.

## Acknowledgments

We would like to thank all the participants who took part in the Dama Health Genetics and Contraception Study for sharing their experiences.

## Data availability statement

The participants of this study did not give written consent for their data to be shared publicly, so due to the sensitive nature of the research supporting data is not available, avoiding identifying any of the authors prior to peer review

